# Is the vitamin D status of patients with COVID-19 associated with reduced mortality?

**DOI:** 10.1101/2021.03.25.21254310

**Authors:** Paulo R Bignardi, Paula de Andrade Castello, Bruno de Matos Aquino, Vinicius Daher Alvares Delfino

## Abstract

**Objective:** A systematic review with meta-analysis was performed to assess a possible association between plasma vitamin D levels and mortality in patients with COVID – 19.

**Methods:** PubMed, EMBASE, and Cochrane Library databases were searched. Studies involving COVID-19 patients that reported an association between plasma vitamin D levels and COVID-19 mortality published until February 5, 2021, were included. The risk ratio (RR) and confidence interval (CI) were pooled using a fixed-effects or random-effects model.

**Results:** A total of 11 studies that measured plasma vitamin D levels at admission were included in the meta-analysis, ten cohorts and one case-controls. Low plasma vitamin D levels (25(OH)D) in patients with COVID-19 were not associated with mortality (RR=1.35, 95%CI 0.84–1.86). Subgroup analysis by vitamin D cut-off (<20 or 25 ng/ml and <10 or 12 ng/ml) showed were not associated with mortality. When the RR in mortality analysis was calculated included four studies that did not perform adjusted analysis for confounding factors, the result was 1.43 (95% CI 1.18-1.69), suggesting that confounders may have led many observational studies to incorrectly estimate the association between vitamin D status and mortality in COVID-19 patients.

**Conclusion:** Deficient vitamin D levels were not associated with a higher mortality rate in patients with COVID-19. Randomized clinical trials are needed to assess this association.

## INTRODUCTION

Since the first case registered in December 2019 in the city of Wuhan, Hubei Province, China, COVID-19 has spread rapidly throughout the world for presenting strong contagious and infectious characteristics [1–3], which have caused 2,309,370 deaths until February 8^th^, 2020, in 198 countries[4].

Worldwide data from the pandemic demonstrate a mortality rate of 0.9% in patients without comorbidities, which increases progressively based on the number of comorbidities and the patients’ age[5]. Studies relating vitamin D levels to Acute respiratory infections[6] led to the carrying out an ecological study that showed that countries where the plasma mean vitamin D in the population are low, had higher infection rates and mortality SARS-CoV2[7].

Isaia et al.[8] found a correlation between regions with higher levels of solar ultraviolet (UV) radiation and lower rates of morbidity and mortality related to COVID-19. The hypothesis discussed by the authors is that it may be related to vitamin D levels. Exposure to UV radiation determines the photo-conversion of the 7-dehydrocholesterol in the skin to pre-vitamin D3[9]. There are two forms of vitamin D (D2 and D3), and the primary source of vitamin D3 (about 80% of the vitamin D organic stores) is via UV[10]. Vitamin D from the skin and diet is metabolized in the liver to 25-hydroxyvitamin D (25(OH)D), the form used to determine patients’ vitamin D status, is hydroxylated in the kidneys in the active form 1,25 dihydroxyvitamin D (1,25(OH)_2_D).

Vitamin D insufficiency is defined as a blood level of 25(OH)D <30 ng/mL, vitamin D deficiency is a blood level of 25(OH)D <20 ng/mL, and severe vitamin D deficiency as a blood level of 25(OH)D < 10 ng/ml [11, 12] or <12 ng/ml, according to some authors[13, 14]. Small observational studies relating vitamin D deficiency or insufficiency to COVID-19 outcomes have emerged with divergent results[15–19]. Thus, we conducted a systematic review followed by meta-analysis to assess a possible association between plasma vitamin D levels and mortality in patients with COVID-19.

## METHODS

### Data Search

Two investigators searched MEDLINE, EMBASE, and Cochrane Library. Studies published until February 5, 2021, were included. The following search strategy was used: (coronavirus OR “coronavirus infections” OR COVID-19 OR “severe acute respiratory syndrome coronavirus 2”) AND (“vitamin D). The language of the searches was limited to English, Spanish, and Portuguese.

### Study selection

Two independent authors screened the studies. Disagreements were resolved through discussion among all authors. Summaries of retrieved articles were reviewed to exclude irrelevant studies, followed by reading full text for screening.

Studies were included if they met the following inclusion criteria: (1) Hospitalized patients with COVID-19. All studies used throat swab SARS-CoV-2 real-time reverse transcription-polymerase chain reaction (RT-PCR) nucleic acid to confirm COVID-19 diagnosis; (2) COVID-19 patients with deficient or insufficient vitamin D status and plasma vitamin D (25(OH)D) levels measured on hospital admission (up to 48 hours after diagnosis); (3) COVID-19 patients with sufficient vitamin D status as the comparator; (4) examined the association between vitamin D status and mortality. Ecological, case reports, cross-sectional, animal model study, and studies that did not mention that plasma vitamin D levels were measured at admission were excluded.

### Data Extraction

Eligible studies included assessing death or disease severity in individuals with plasma vitamin D. They should provide the odds ratio (OR), risk ratio (RR), or hazard ratio (HR) with 95% confidence intervals (CI). Inclusion was not restricted by study size.

Data extracted included authors, study design, country of origin, demographic characteristics (age and sample size), COVID-19 diagnosis, outcomes relevant studies on the study question. Two independent investigators performed data extraction. Disagreements were resolved through discussion among all authors.

## Results evaluation

The analysis focused on the mortality outcome in patients with COVID-19 within the low plasma vitamin D levels group compared with the sufficient plasma vitamin D level group. We performed one stratified analysis by cut-off vitamin D (<20 or 25 ng/ml and <10 or 12 ng/ml), and a second stratified analysis using studies that provided analysis adjusted for confounding factors and studies that did not. For the present meta-analysis, vitamin D levels were standardized to ng/ml; studies that provided vitamin D levels in nmol/l had these levels transformed into ng/ml[10]. Besides, a sensitivity analysis was performed, omitting each study to detect the influence on the overall effect’s estimate.

### Quality assessment and statistical analysis

This systematic review was conducted following the Preferred Items guidelines for Reporting for Systematic Reviews and Meta-Analysis (PRISMA), and this study has not been registered.

The New-Castle-Ottawa quality scale[20] was used to evaluate the quality of the observational studies. Studies included in the meta-analysis reported OR, HR, or RR. For studies that did not report these effects, the RR calculation was based on the Cochrane Handbook for Systematic Reviews [21].

Effect estimates with the most significant degree of adjustment for potential confounding factors were extracted. HR was considered comparable to RR. For studies that reported OR, a corrected RR was computed as already described[22]. Pooled RR and 95% confidence interval (CI) were calculated using a fixed or random-effects model according to the studies homogeneity. The Cochran Q test and the I^2^ statistic were used to evaluate the statistical significance and degree of heterogeneity between the studies, respectively. The statistic I^2^≥50% reveals substantial heterogeneity. Finally, the publication bias was examined by the Egger test and a funnel plot. All analyses were performed with Stata/SE v.14.1 software (StataCorpLP, USA).

## RESULTS

### Characteristics of the selected studies

Seven-hundred and seventy-two (772) studies were identified through database research. Of these, 653 studies were duplicate articles or were excluded based on predetermined eligibility criteria during title/abstract review. According to the inclusion and exclusion criteria, we identified 11 studies[23, 24, 33, 25–32] that were eligible for this review and meta-analysis (Fig. 1), involving 1,347 participants. Ten were cohort[23, 24, 26–28, 30–33], and one case-control[25]. The characteristics of selected studies and participants are summarised in Table 1.

**Table 1.**
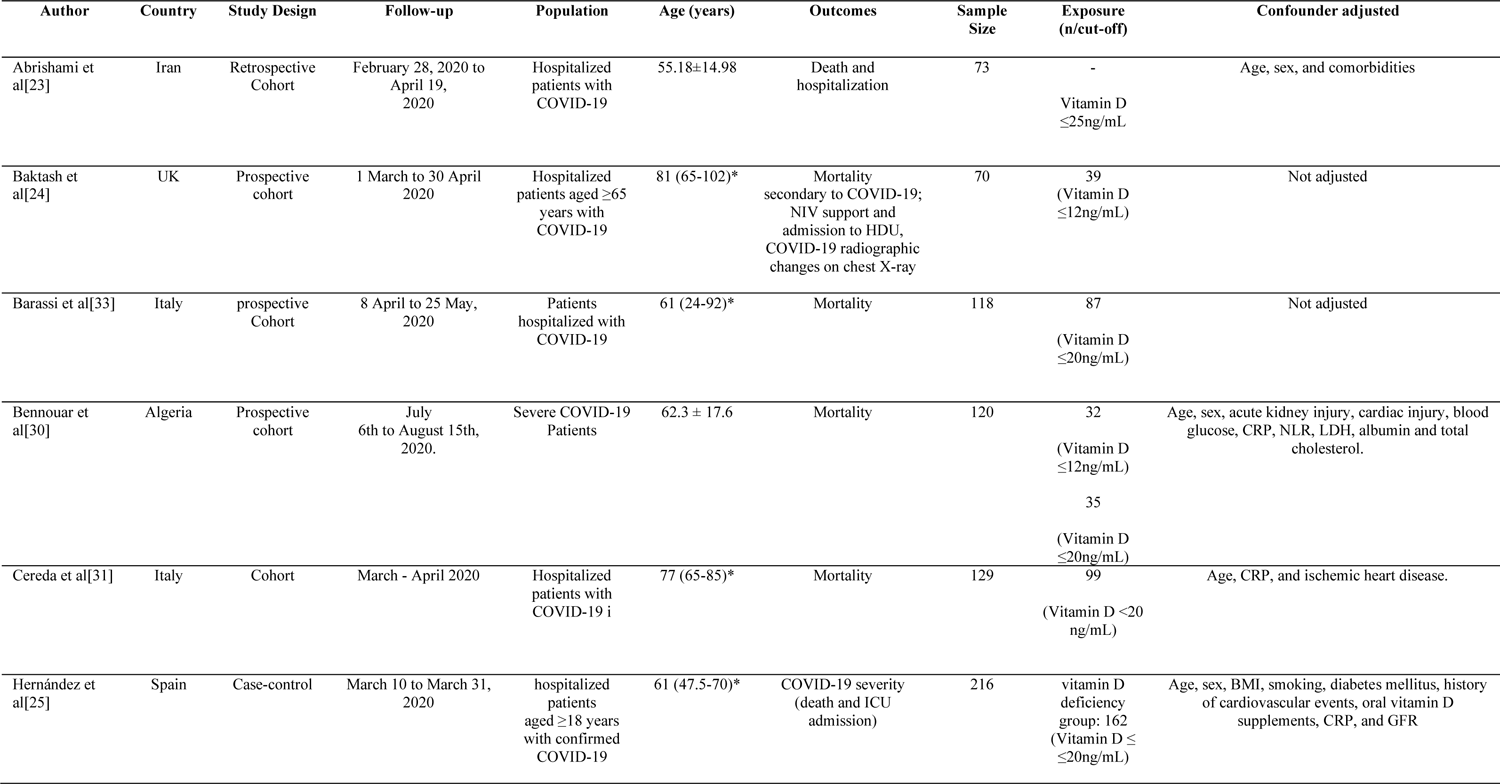
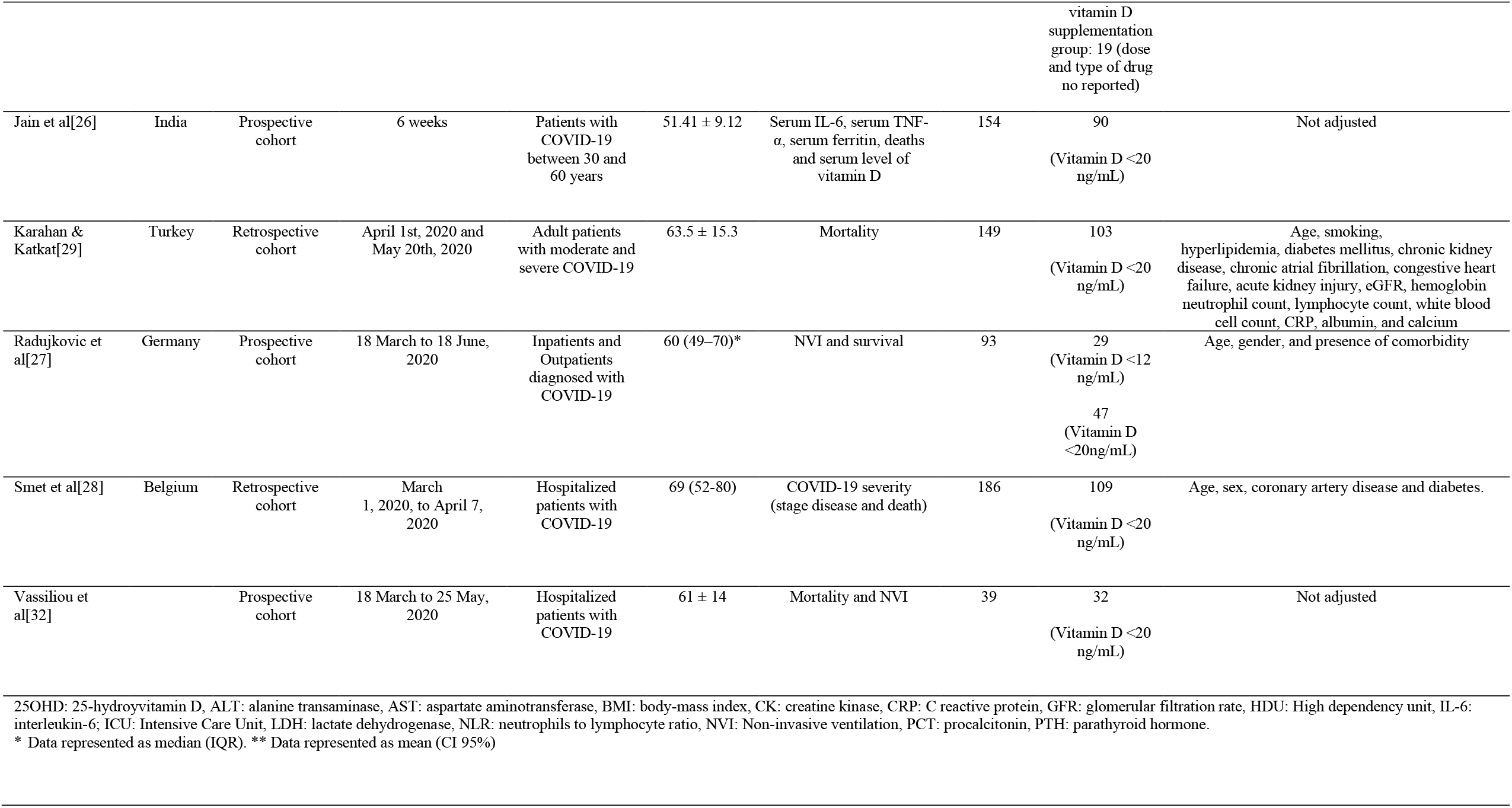
Characteristics of studies selected.

| Author | Country | Study Design | Follow-up | Population | Age (years) | Outcomes | Sample Size | Exposure (n/cut-off) | Confounder adjusted |
| --- | --- | --- | --- | --- | --- | --- | --- | --- | --- |
| Abrishami et al[23] | Iran | Retrospective Cohort | February 28, 2020 to April 19, 2020 | Hospitalized patients with COVID-19 | 55.18±14.98 | Death and hospitalization | 73 | -<br>Vitamin D ≤25ng/mL | Age, sex, and comorbidities |
| Baktash et al[24] | UK | Prospective cohort | 1 March to 30 April 2020 | Hospitalized patients aged ≥65 years with COVID-19 | 81 (65-102)* | Mortality secondary to COVID-19; NIV support and admission to HDU, COVID-19 radiographic changes on chest X-ray | 70 | 39 (Vitamin D ≤12ng/mL) | Not adjusted |
| Barassi et al[33] | Italy | prospective Cohort | 8 April to 25 May, 2020 | Patients hospitalized with COVID-19 | 61 (24-92)* | Mortality | 118 | 87 (Vitamin D ≤20ng/mL) | Not adjusted |
| Bennouar et al[30] | Algeria | Prospective cohort | July 6th to August 15th, 2020. | Severe COVID-19 Patients | 62.3 ± 17.6 | Mortality | 120 | 32 (Vitamin D ≤12ng/mL)<br>35 (Vitamin D ≤20ng/mL) | Age, sex, acute kidney injury, cardiac injury, blood glucose, CRP, NLR, LDH, albumin and total cholesterol. |
| Cereda et al[31] | Italy | Cohort | March - April 2020 | Hospitalized patients with COVID-19 i | 77 (65-85)* | Mortality | 129 | 99 (Vitamin D <20 ng/mL) | Age, CRP, and ischemic heart disease. |
| Hernández et al[25] | Spain | Case-control | March 10 to March 31, 2020 | hospitalized patients aged ≥18 years with confirmed COVID-19 | 61 (47.5-70)* | COVID-19 severity (death and ICU admission) | 216 | vitamin D deficiency group: 162 (Vitamin D ≤20ng/mL) | Age, sex, BMI, smoking, diabetes mellitus, history of cardiovascular events, oral vitamin D supplements, CRP, and GFR |

|  |  |  |  |  |  |  |  | vitamin D<br>supplementation<br>group: 19 (dose<br>and type of drug<br>no reported) |  |
| --- | --- | --- | --- | --- | --- | --- | --- | --- | --- |
| Jain et al[26] | India | Prospective<br>cohort | 6 weeks | Patients with<br>COVID-19<br>between 30 and<br>60 years | 51.41 ± 9.12 | Serum IL-6, serum TNF-α, serum ferritin, deaths<br>and serum level of<br>vitamin D | 154 | 90<br>(Vitamin D <20<br>ng/mL) | Not adjusted |
| Karahan &<br>Katkat[29] | Turkey | Retrospective<br>cohort | April 1st, 2020 and<br>May 20th, 2020 | Adult patients<br>with moderate and<br>severe COVID-19 | 63.5 ± 15.3 | Mortality | 149 | 103<br>(Vitamin D <20<br>ng/mL) | Age, smoking,<br>hyperlipidemia, diabetes mellitus, chronic kidney<br>disease, chronic atrial fibrillation, congestive heart<br>failure, acute kidney injury, eGFR, hemoglobin<br>neutrophil count, lymphocyte count, white blood<br>cell count, CRP, albumin, and calcium |
| Radujkovic et<br>al[27] | Germany | Prospective<br>cohort | 18 March to 18 June,<br>2020 | Inpatients and<br>Outpatients<br>diagnosed with<br>COVID-19 | 60 (49–70)* | NVI and survival | 93 | 29<br>(Vitamin D <12<br>ng/mL)<br>47<br>(Vitamin D<br><20ng/mL) | Age, gender, and presence of comorbidity |
| Smet et al[28] | Belgium | Retrospective<br>cohort | March<br>1, 2020, to April 7,<br>2020 | Hospitalized<br>patients with<br>COVID-19 | 69 (52-80) | COVID-19 severity<br>(stage disease and death) | 186 | 109<br>(Vitamin D <20<br>ng/mL) | Age, sex, coronary artery disease and diabetes. |
| Vassiliou et<br>al[32] |  | Prospective<br>cohort | 18 March to 25 May,<br>2020 | Hospitalized<br>patients with<br>COVID-19 | 61 ± 14 | Mortality and NVI | 39 | 32<br>(Vitamin D <20<br>ng/mL) | Not adjusted |

25OHD: 25-hydroxyvitamin D, ALT: alanine transaminase, AST: aspartate aminotransferase, BMI: body-mass index, CK: creatine kinase, CRP: C reactive protein, GFR: glomerular filtration rate, HDU: High dependency unit, IL-6: interleukin-6; ICU: Intensive Care Unit, LDH: lactate dehydrogenase, NLR: neutrophils to lymphocyte ratio, NVI: Non-invasive ventilation, PCT: procalcitonin, PTH: parathyroid hormone.
\* Data represented as median (IQR). \*\* Data represented as mean (CI 95%)

**Fig. 1.**
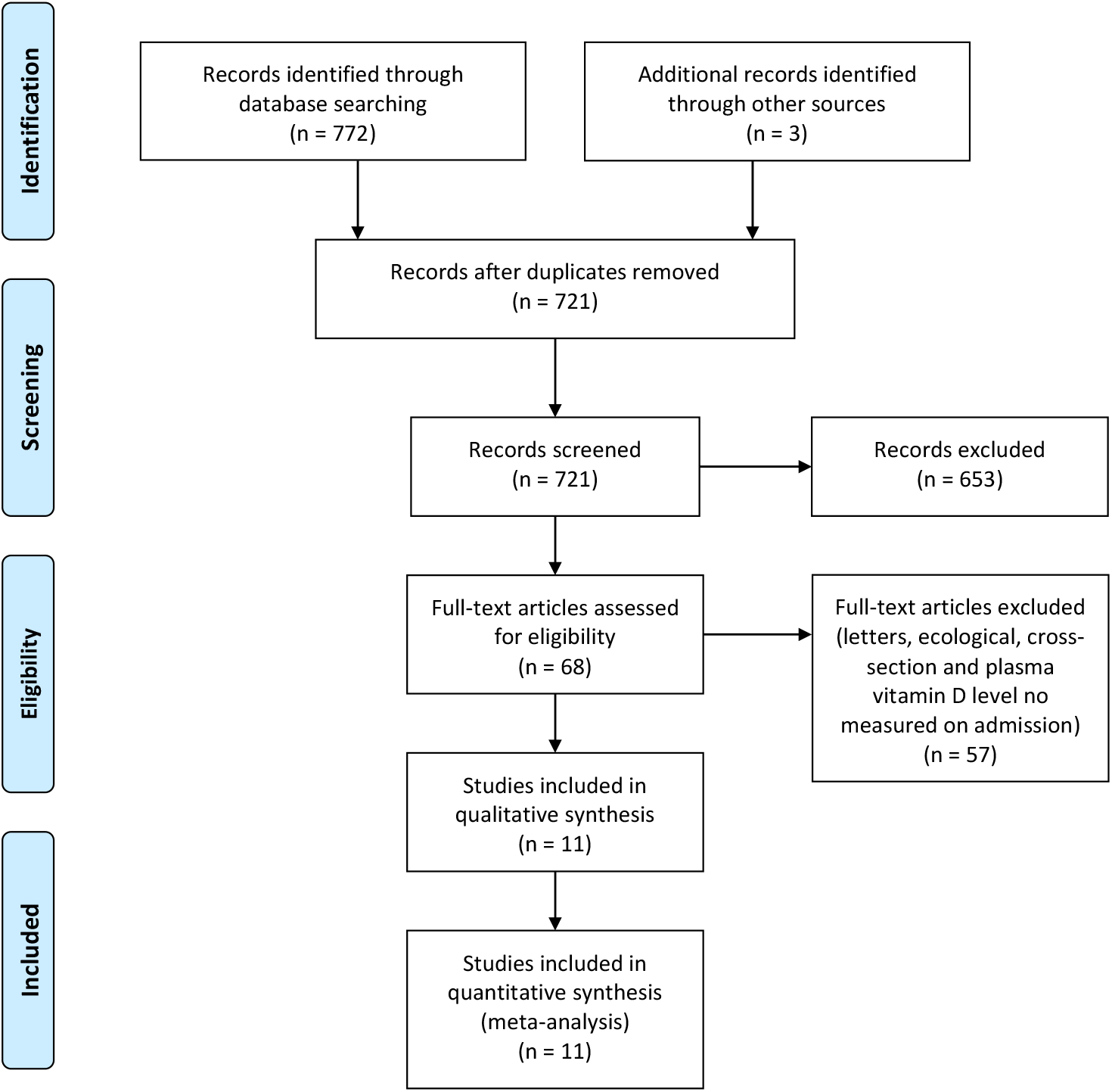
Flow chart of study selection

### Plasma vitamin D level and mortality in patients with COVID-19

The mortality outcome was extracted from the 11 studies. Bennouar et al.[29] and Radujkovic et al.[26] were included twice because they analysed two cut-offs of vitamin D, <10 ng/ml and <20 ng/ml. Figure 2 shows that the mortality in patients with deficient plasma vitamin D levels did not differ from patients with sufficient plasma vitamin D levels (RR=1.35, 95%CI 0.84–1.86, I^2^=62.3%). Analysis by vitamin D level (Fig. 2) showed a RR=1.34 (95% CI 0.79-1.89, I2 = 69.6%) for studies with plasma vitamin D <20 or 25 ng/ml, and RR=1.50 (95% CI 0.02-3.02, I2 = 2.4%) for studies with plasma vitamin D <10 or 12 ng/ml.

**Fig. 2.**
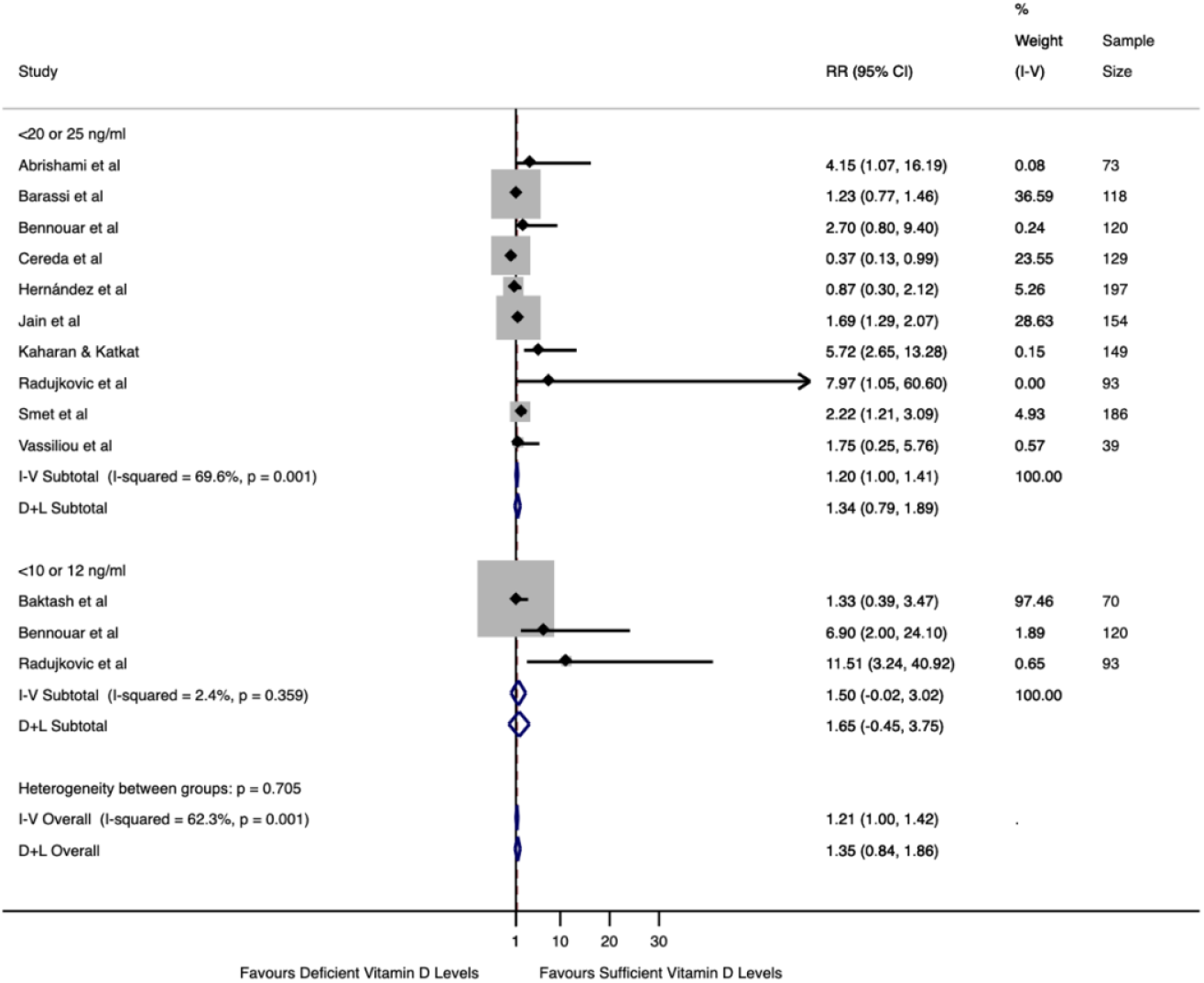
Association of plasma vitamin D levels with mortality in COVID-19 patients by cut-off of 25(OH)D.

Subgroup analysis (Fig. 3), including studies that performed adjustments for age and at least one more confounding factor (obesity, hypertension, diabetes, chronic kidney disease, and cardiovascular disease), revealed an RR=1.49 (95% CI 0.44-2.55, I^2^ = 60.1%). The analysis, including studies that did not mention adjustment for confounders, showed an increased mortality risk (RR=1.43, 95%CI 1.18-1.69, I^2^=2.2%).

**Fig. 3.**
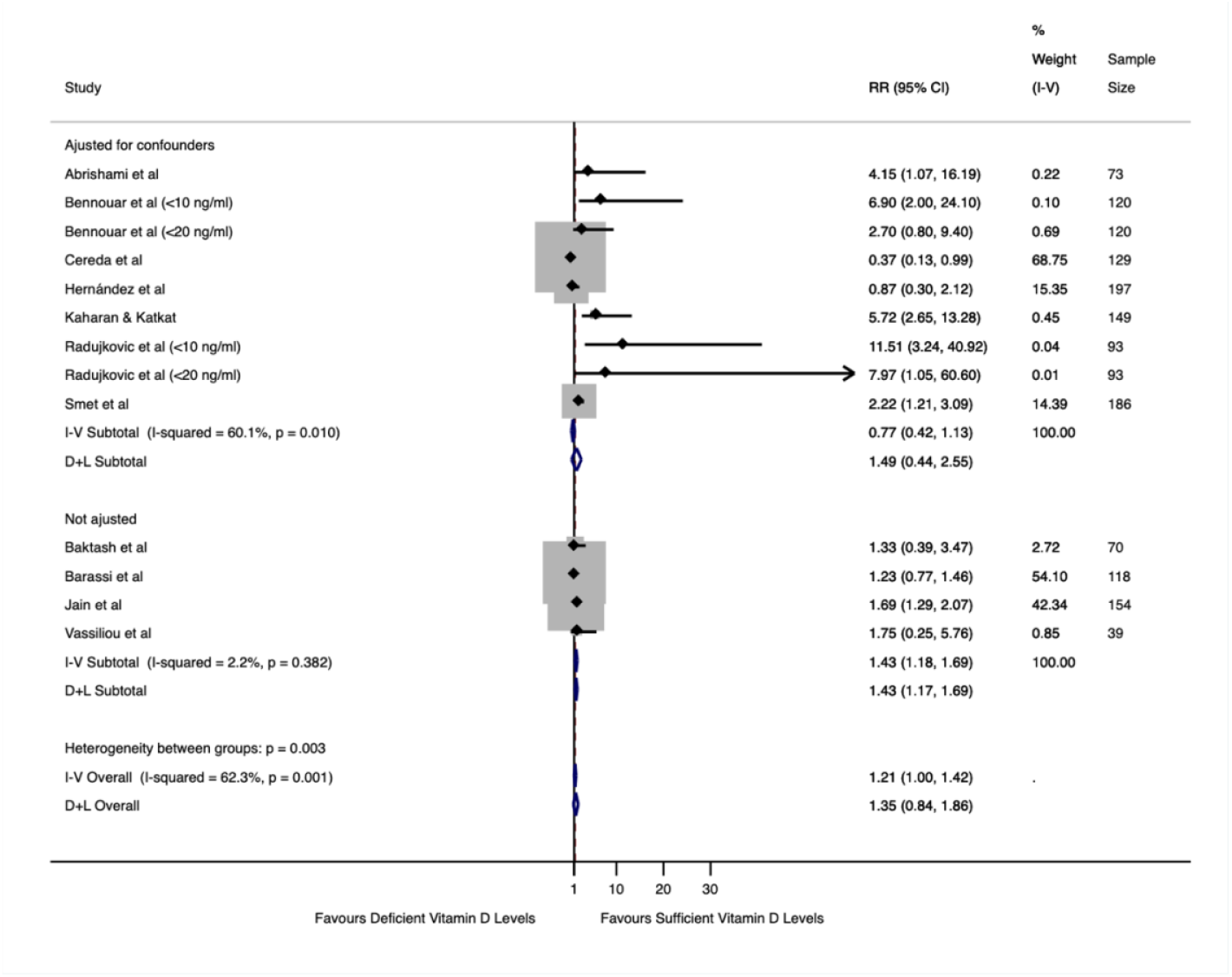
Association between plasma vitamin D levels and mortality in patients with COVID-19, including studies that adjusted the analysis for age and at least one more confounder (age, obesity, hypertension, diabetes, kidney disease and Cardiovascular Disease) and studies without adjust for confounders.

### Sensitivity analyses, assessment of heterogeneity and risk of bias

Table 2 shows sensitivity analyses performed, excluding one study that adjusted confounding factors at a time for the mortality outcome. There was no change in Cochran’s Q test, and the I^2^ varied from 24.1% to 65.0%, persisting the same results in all scenarios.

**Table 2.** Sensitive analysis for the mortality outcome, including studies that performed adjusted analysis for confounders.

| **Study omitted** | **RR** | **95% CI** | **I<sup>2</sup>** | **P-value\*** |
| --- | --- | --- | --- | --- |
| Abrishami et al | 1.45 | 0.38-2.53 | 63.7% | >0.05 |
| Bennouar et al (<10 ng/ml) | 1.49 | 0.39-2.50 | 62.9% | >0.05 |
| Bennouar et al (<20 ng/ml) | 1.44 | 0.34-2.54 | 63.7% | >0.05 |
| Cereda et al | 1.89 | 0.84-2.94 | 24.1% | >0.05 |
| Hernández et al | 2.05 | 0.45-3.65 | 65.0% | >0.05 |
| Kaharan & Katkat | 1.30 | 0.31-2.29 | 58.1% | >0.05 |
| Radujkovic et al (<12 ng/ml) | 1.46 | 0.41-2.50 | 62.7% | >0.05 |
| Radujkovic et al (<20 ng/ml) | 1.51 | 0.43-2.58 | 64.7% | >0.05 |
| Smet et al | 0.88 | 0.04-1.72 | 25.6% | >0.05 |

\* value for heterogeneity among studies assessed with Cochran's Q test.

The risk of bias in studies is detailed in Table 3. The estimated bias coefficient w as 0.103, with *P*-value >0.05, indicating no small-study effects. A funnel plot was performed but failed to detect possible small study effects (Fig. 4). Therefore, the tests provide weak evidence for the presence of publication bias.

**Table 3.** Risk of bias from studies included in meta-analyzes. (Newcastle Ottawa scale)

| **CASE-CONTROL STUDIES** | **SELECTION** |  |  |  | **COMPARABILITY** |  | **EXPOSURE** |  |  | **TOTAL OF STARS** |
| --- | --- | --- | --- | --- | --- | --- | --- | --- | --- | --- |
|  | 1 | 2 | 3 | 4 | 1A | 1B | 1 | 2 | 3 |  |
| Hernández et al | Y | Y | Y | Y | Y | Y | N | Y | Y | 8 |
| **COHORT STUDIES** | **SELECTION** |  |  |  | **COMPARABILITY** |  | **OUTCOME** |  |  | **TOTAL OF STARS** |
|  | 1 | 2 | 3 | 4 | 1A | 1B | 1 | 2 | 3 |  |
| Abrishami et al | Y | Y | Y | Y | Y | Y | Y | Y | Y | 9 |
| Baktash et al | Y | Y | Y | Y | Y | N | Y | Y | Y | 8 |
| Barassi et al | Y | Y | Y | Y | Y | N | Y | Y | Y | 8 |
| Bennouar et al | Y | Y | Y | Y | Y | Y | Y | Y | Y | 9 |
| Cereda et al | Y | Y | Y | Y | Y | Y | Y | Y | Y | 9 |
| Jain et al | Y | Y | Y | Y | N | N | Y | Y | Y | 7 |
| Karahan & Katkat | Y | Y | Y | Y | Y | Y | Y | Y | Y | 9 |
| Radujkovic et al | Y | Y | Y | Y | Y | Y | Y | Y | Y | 9 |
| Smet et al | Y | Y | Y | Y | Y | Y | Y | Y | Y | 9 |
| Vassiliou et al | N | Y | Y | Y | Y | N | Y | Y | Y | 7 |

**Fig. 4.**
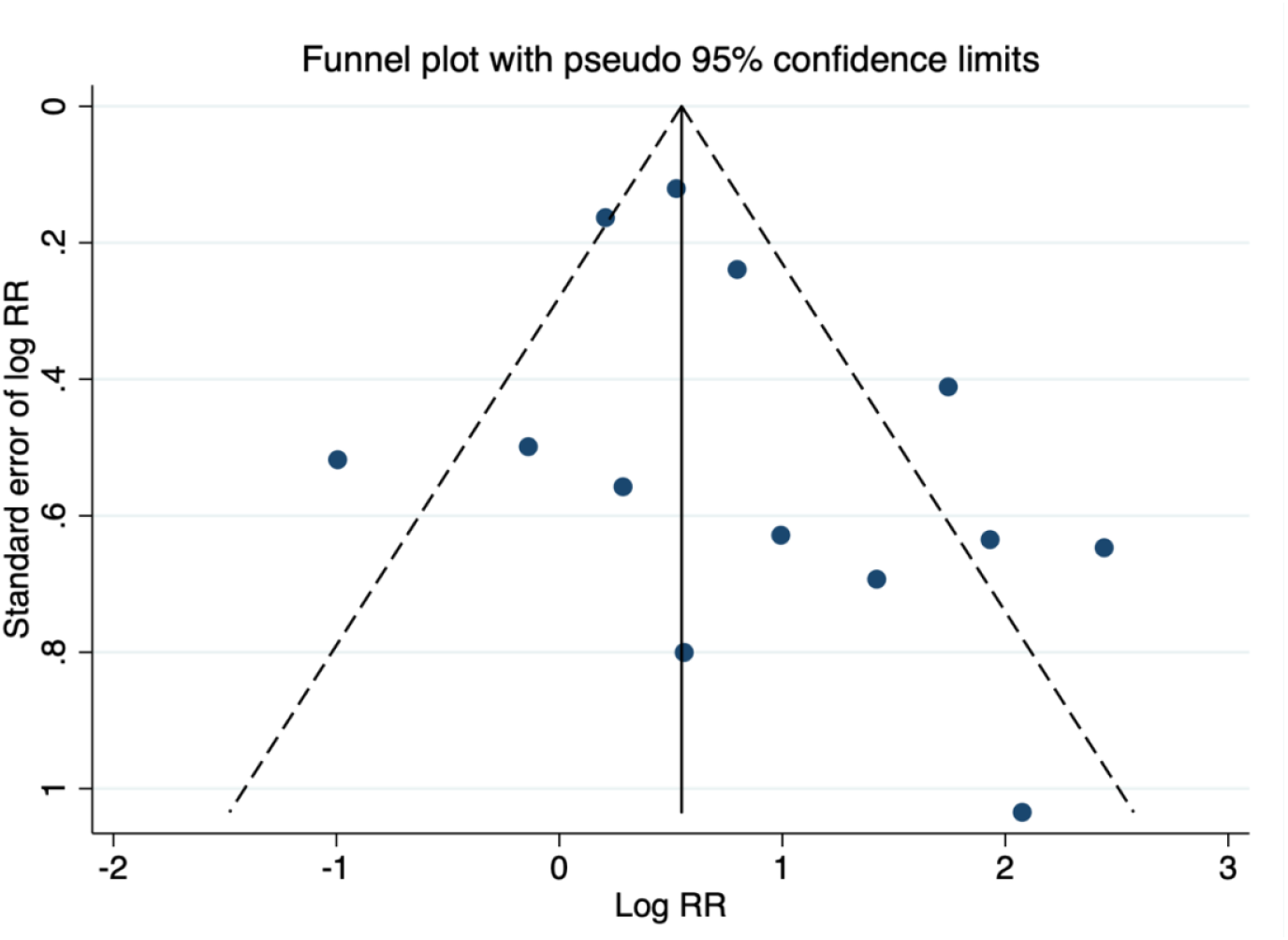
Funnel plot, using data from 11 studies associating plasma vitamin D levels and mortality. Two studies were included twice because they analysed two cut-offs of 25(OH)D.

## DISCUSSION

This is the first systematic review with meta-analysis to assess only studies that measured plasma vitamin D levels in patients close to the diagnosis of COVID-19. Observational studies have associated low plasma vitamin D levels and poor outcomes in patients with COVID-19. However, many of these studies used the plasma vitamin D level measured from a few months to years before the diagnosis of COVID-19[34, 35]. The biological variability of vitamin D is known, and its plasma levels can vary from 13 to 26% in 4 months. Plasma vitamin D can also vary with age and the appearance of comorbidities[36–40].

This study did not show an association between vitamin D status in COVID-19 patients and increased mortality risk. The overall analysis of mortality did not reveal an association between low plasma vitamin D levels and mortality. Likewise, the subgroup analysis with the studies that perform adjusted analysis with age and at least one more confounder did not show this association.

However, an analysis including only studies that did not performed adjusted analysis with confounders revealed an association between low plasma vitamin D levels and an increased risk of death (1.43-fold), which shows that confounding factors may have driven many previous studies results. Therefore, our meta-analysis suggests that vitamin D status does not have a causal effect on mortality in COVID-19 patients.

Patchen et al. aiming to investigate the causality of the association between serum vitamin D status and severity of COVID-19 infection, studied single nucleotide polymorphisms (SNPs) related to the risk of vitamin D deficiency. They found no association between genetically predicted differences in long-term vitamin D nutritional status and poor outcomes in patients with COVID-19[41].

Hypovitaminosis D shares many risk factors with the severe form of COVID-19. Older age, obesity, chronic kidney disease, diabetes, hypertension, and cardiovascular disease are some critical risk factors that have been reported as associated with the severity of COVID-19[42–45]. There is evidence that vitamin D deficiency can be caused by older age, obesity, and chronic kidney disease [46–51]. Type 2 diabetes, hypertension, and cardiovascular disease also are associated with vitamin D deficiency [52, 53].

It is known that vitamin D has a role in modulating the immune response, and its deficiency is associated with an increased risk of developing viral and bacterial infections [54, 55]. However, it may be that its role in preventing the mortality of patients with COVID-19 is outweighed by other mechanisms involved in the pathophysiology of the disease.

The liver hydroxylates vitamin D to 25(OH)D (Calcifediol), a form found in the blood with a 3-week plasma half-life (whole-body half-life is of 2–3 months)[56]. Plasma levels of Calcifediol are generally used to check people’s vitamin D status. Calcifediol is hydroxylated to its active form 1,25(OH) _2_D, which has a plasma half-life of 4 hours, mainly in the kidneys but also in some other extra-renal sites, including pulmonary epithelial cells[10, 56]. Plasma level of 1,25(OH) _2_D is roughly 1000-fold lower compared with Calcifediol[57].

The kidney is the main organ for the regulation of serum 1,25 (OH) _2_D[58]. It is known that the ACE2 protein, present in renal and lung cells, is a target for SARS-CoV2 to enter these cells[59, 60]. Renal dysfunction caused directly by the virus or indirectly by the presence of acute kidney associated with COVID-19 infection may lead to a reduction in the activity of 1-alpha hydroxylase, the enzyme that converts Calcifediol to 1,25(OH)_2_D[61, 62].

Therefore, a possible hypothesis that could explain our findings would be that tissue damage in the kidneys and to a lesser extent in the lung caused by SARS-CoV2 may lead to an active decrease of 1,25(OH)D, responsible for the majority of biological actions of Vitamin D, but not in the form usually determined in plasma, Calcifediol. Previous studies showed that blood 25(OH)D levels may not correlate with blood 1,25(OH)_2_D levels in some clinical conditions [63–65].

Reduced calcium and phosphorus were found in critically ill COVID-19 patients, which may indicate a reduction of 1,25(OH) _2_D in these patients since the active form of vitamin D is an essential regulator of calcium and phosphorus levels acting in intestinal absorption and renal reabsorption[58].

Despite the well-known autocrine and paracrine production of 1,25(OH) _2_D cells in the immune system, it is difficult to predict whether plasma levels do not influence the immune response against SARS-CoV2. Furthermore, Playford et al. findings that plasma levels of 1,25(OH) _2_D, but not 25(OH)D were associated with cardiovascular risk factors, a common outcome in patients with severe COVID-19[66]. Likewise, Nguyen et al. findings that plasma levels of 1,25(OH)_2_D were a better predictor of mortality by sepsis than Calcifediol[67]. In addition, 1,25(OH)_2_D antiviral action against SARS-CoV2 and other virus has been proposed[68, 69].

This study has several strengths. To our knowledge, this is the first meta-analysis associating plasma vitamin D levels measured on or near hospital admission of patients with COVID-19, and that included studies that performed analysis adjusted with confounders. The main study limitations were analysis that has used different cut-offs of plasma vitamin D, substantial heterogeneity, and observational design of the selected studies.

## CONCLUSION

These results suggest that plasma vitamin D status is not associated with mortality in COVID-19 patients and that confounders may have led to the detrimental effects of low plasma 25(OH)D levels on COVID-19 patients observed in some previous studies. Large randomized clinical trials are needed to assess the effects of vitamin D levels, including 1,25(OH) _2_D, on mortality of hospitalized patients with COVID-19.

## Data Availability

Not applicable

